# Pediatric household transmission of SARS-CoV-2 infection

**DOI:** 10.1101/2021.03.29.21254565

**Authors:** Lauren A. Paul, Nick Daneman, Kevin L. Schwartz, Michelle Science, Kevin A. Brown, Michael Whelan, Ellen Chan, Sarah A. Buchan

**Affiliations:** Health Protection, Public Health Ontario, 661 University Ave., Floor 17, Toronto, ON, M5G 1M1, Canada; Sunnybrook Research Institute, Sunnybrook Health Sciences Centre, 2075 Bayview Ave., Toronto, ON, M4N 3M5, Canada; Division of Infectious Diseases, Sunnybrook Health Sciences Centre, 2075 Bayview Ave., Room B1 03, Toronto, ON, M4N 3M5, Canada; Department of Medicine, University of Toronto, 6 Queen’s Park Cres. W, Floor 3, Toronto, ON, M5S 3H2, Canada; Institute of Health Policy, Management and Evaluation, University of Toronto, 155 College St., Suite 425, Toronto, ON, M5T 3M6, Canada; Dalla Lana School of Public Health, University of Toronto, 155 College St., Room 500, Toronto, ON, M5T 3M7,Canada; Unity Health Toronto – St. Joseph’s Health Centre, 30 The Queensway, Toronto, ON, M6R 1B5, Canada; Division of Infectious Diseases, The Hospital for Sick Children, 555 University Ave., Clinic 7, Toronto, ON, M5G 1X8, Canada; Department of Paediatrics, University of Toronto, 555 University Ave., Black Wing Room 1436, Toronto, ON, M5G 1X8, Canada

**Keywords:** COVID-19, SARS-CoV-2, household, transmission, children, pediatric, age, school-aged

## Abstract

**BACKGROUND:** As a result of low numbers of pediatric cases early in the COVID-19 pandemic, pediatric household transmission of SARS-CoV-2 remains an understudied topic. This study sought to determine whether there are differences in the odds of household transmission for younger children compared to older children.

**METHODS:** We assembled a cohort of all individuals in Ontario, Canada with laboratory-confirmed SARS-CoV-2 infection between June 1 and December 31, 2020. The cohort was restricted to individuals residing in private households (N=132,232 cases in 89,191 households), identified through an address matching algorithm. Analysis focused on households in which the index case was aged <18 years. Logistic regression models were fit to estimate the association between age group of pediatric index cases (0-3, 4-8, 9-13, and 14-17 years) and odds of household transmission.

**RESULTS:** A total of 6,280 households had pediatric index cases, and 1,717 (27.3%) experienced secondary transmission. Children aged 0-3 years had the highest odds of household transmission compared to children aged 14-17 years (model adjusted for gender, month of disease onset, testing delay, and average family size: 1.43, 95% CI: 1.17-1.75). This association was similarly observed in sensitivity analyses defining secondary cases as 2-14 days or 4-14 days after the index case, and stratified analyses by presence of symptoms, association with a school/childcare outbreak, or school/childcare reopening. Children aged 4-8 years and 9-13 years also had increased odds of transmission (4-8: 1.40, 95% CI: 1.18-1.67; 9-13: 1.13, 95% CI: 0.97-1.32).

**CONCLUSIONS:** This study suggests that younger children are more likely to transmit SARS-CoV-2 infection compared to older children, and the highest odds of transmission was observed for children aged 0-3 years. Differential infectivity of pediatric age groups has implications for infection prevention controls within households, as well as schools/childcare, to minimize risk of household secondary transmission.

## Introduction

The role of children in the transmission of severe acute respiratory syndrome coronavirus 2 (SARS-CoV-2) infection requires further study. Early in the pandemic, when countries implemented lockdown measures, close contact was mostly limited to households and testing strategies tended to prioritize healthcare workers and symptomatic individuals.^1–4^ As a result there were relatively few diagnosed pediatric cases of coronavirus disease 2019 (COVID-19)^5^ and it appeared that the proportion of children involved in the transmission of infection was small.^2,6,7^ Since many jurisdictions relaxed public health measures and re-opened educational facilities in the fall of 2020, the number of pediatric COVID-19 cases has grown, providing the opportunity to better characterize the infectivity of children.

Household studies to date have typically only compared infectivity between young and old individuals, often grouping children with young adults^8–12^ or dichotomizing age to older adults versus younger adults/children.^13,14^ These studies have reported mixed results, with some finding that older age (_≥_20 years) was associated with increased transmission^6,8–10,15,16^, one study finding younger age (<20 years)^12^, and others finding no age effect.^11,13– 15,17–19^ Conversely, few household studies have examined differences in transmission among children^15,16,20–23^, likely due to insufficient sample size.^2,24^ Two meta-analyses reported no significant differences between younger children and older children for household susceptibility to SARS-CoV-2^2,15^, however it remains unclear if this holds true for infectivity. These findings warrant a closer look at household transmission of SARS-CoV-2 by children, and whether there are any differences in the likelihood of transmission for particular age groups.

We sought to conduct an age analysis of residents aged 0-17 years in Ontario, Canada that were the index case of SARS-CoV-2 infection in their household between June and December 2020. Pediatric index cases were divided into four age groups (0-3, 4-8, 9-13, and 14-17 years) to provide a more granular picture of any age differences. We were also interested in comparing characteristics of index cases by age group, exploring the direction of transmission by age, and assessing whether factors such as symptoms, school/childcare reopening, or school/childcare outbreaks were associated with differences in the odds of transmission from children to their household members.

## Methods

### Study population

We derived the study cohort from case data that was reported in provincial disease systems by health units across Ontario, Canada’s most populous province. All individuals with laboratory-confirmed SARS-CoV-2 infection between June 1 and December 31, 2020 were included. We obtained ethics approval from Public Health Ontario’s Research Ethics Board.

### Identification of private households

Addresses of all cases were reviewed and classified as either private households, defined as individual houses or apartments/suites within multi-unit dwellings, or congregate settings (e.g., homeless shelters or long-term care homes). We excluded any individuals with missing or incomplete address information, individuals residing in congregate settings, and individuals identified as residing in multi-unit dwellings but missing suite information. Addresses were then matched between cases using a natural language processing algorithm from Python’s “sklearn” library in order to identify multi-case households. Details of the address matching process have been described previously.^25^

### Outcomes

The outcome of interest was secondary household transmission of SARS-CoV-2 infection by a pediatric index case (aged 0-17 years). Index cases were defined as the earliest case of a household, and were identified by comparing symptom onset dates of all cases in the household.^2^ If symptom onset date was missing we used specimen collection date as a proxy; no cases were missing both dates. Secondary cases were defined as cases (adult or pediatric) that had disease onset 1-14 days after the index case, as per previous studies of household transmission.^2,21,22,25^ We excluded households with index cases missing age (N=12), as well as households with multiple index cases (i.e., multiple cases occurring on the earliest case date of the household; N=4,335), as they would present challenges for estimating associations between household transmission and characteristics of the index case.

### Individual-level and neighbourhood-level characteristics of index cases

The main exposure of interest was age group of the index case: 0-3, 4-8, 9-13, and 14-17 years. We also a priori selected a group of individual-level and neighborhood-level characteristics of the index case to adjust for in the models. At the individual-level, we included gender, month of disease onset, and testing delay between symptom onset and specimen collection. For the testing delay, we additionally categorized cases that were asymptomatic, identified as cases that were missing symptom onset date (thus specimen collection date was used) and were reported as asymptomatic in provincial reportable disease systems. As we only had individual-level household size information reported for 59.6% of index cases, we also included average family size for the neighbourhood of the index case. Average family size was available from 2016 Canadian census records for aggregate dissemination areas across Ontario, representing areas of approximately 5,000-15,000 persons.

For stratified and sensitivity analyses, we additionally included information on the presence of symptoms, whether the index case was linked to a school/childcare outbreak, and whether the index case’s disease onset was before or after school/childcare reopening (depending on the age of the index case). In Ontario, schools reopened in mid-September 2020 and childcare reopened mid-June 2020.

### Statistical analysis

We carried out descriptive analyses to assess the characteristics of pediatric index cases across the four age groups, and included index cases aged >17 years for comparison. Direction of transmission from index case to secondary case by age was also examined. We then applied three logistic regression models to obtain odds ratios (OR) and 95% confidence intervals (CI) for the associations between index case age group and odds of household transmission: (1) a crude, unadjusted model; (2) a model adjusted for gender and month of disease onset (adjusted model 1); and (3) a model adjusted for gender, month of disease onset, testing delay, and average family size (adjusted model 2).

For stratified analyses, these models were re-fit to subsets of the data broken down by three additional index case characteristics: (1) presence of symptoms; (2) association with a school/childcare outbreak; and (3) disease onset before school/childcare reopening.

For sensitivity analyses, we first updated the definition of secondary cases to increase certainty of the direction of transmission from the index case; we re-ran the analyses with secondary cases that developed symptoms 2-14 days after the index case and 4-14 days after the index case. Second, we adjusted for individual-level household size instead of neighbourhood-level average family size in the subset of index cases that had this information available. Third, we ran the analysis on a symptomatic cohort, restricting to index cases and secondary cases that had symptoms reported or a symptom onset date available. Last, we re-ran the symptomatic cohort analysis with the adjusted definitions for secondary cases (i.e., 2-14 days and 4-14 days).

## Results

Between June and December 2020, a total of 6,280 private households had a pediatric index case (Figure 1). The mean age of pediatric index cases was 10.7 years and 45.6% were female. Of the 6,280 households, 1,717 (27.3%) experienced secondary household transmission, leading to a median of 2 secondary cases (percentiles: 25^th^=1 case, 75^th^=2 cases, 90^th^=3 cases). Pediatric index cases most frequently transmitted infection to individuals aged 0-20 years or 30-50 years, with older children tending to transmit to older individuals in those age ranges (Figure 2).

**Figure 1.**
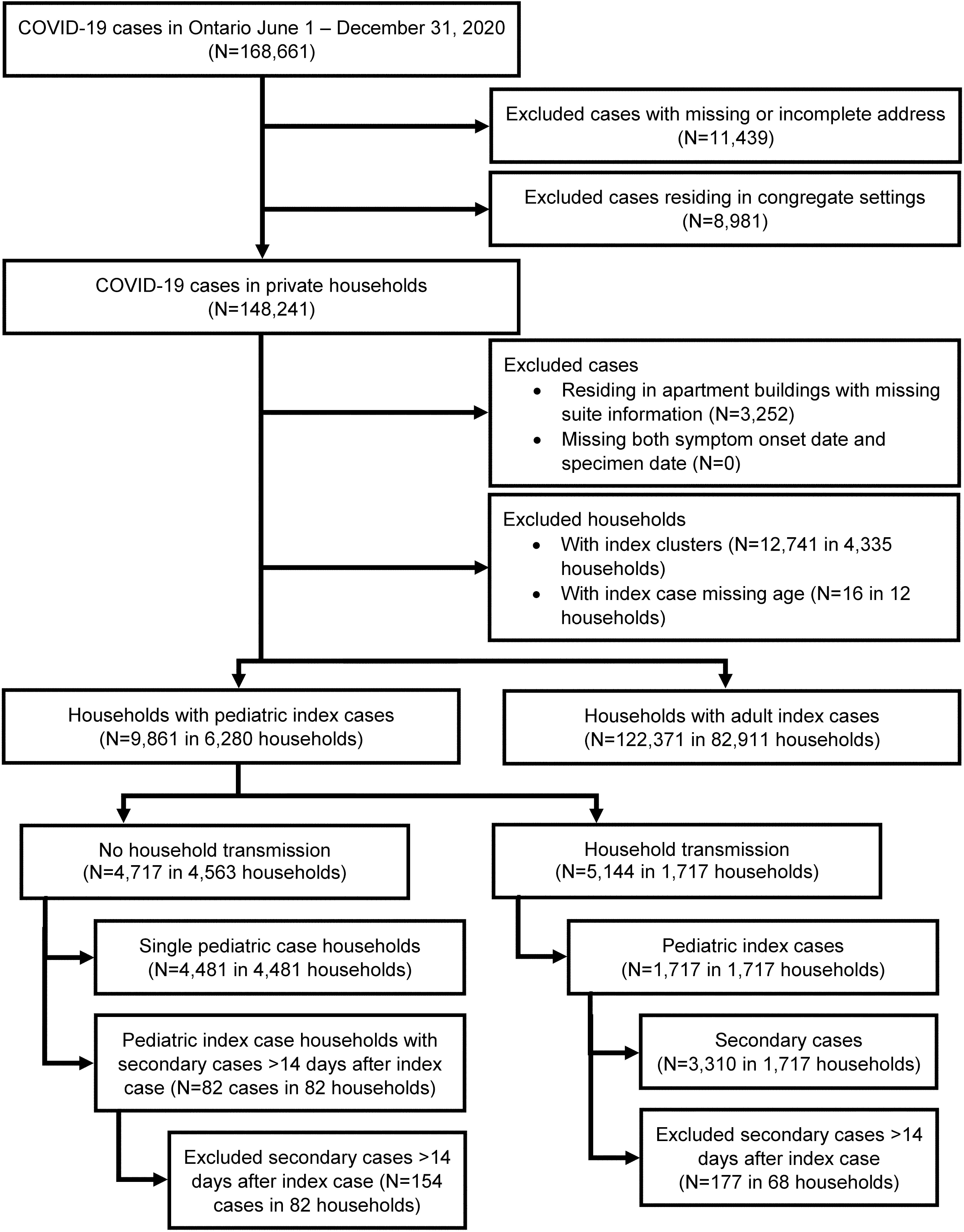
Flow diagram of study cohort

**Figure 2.**
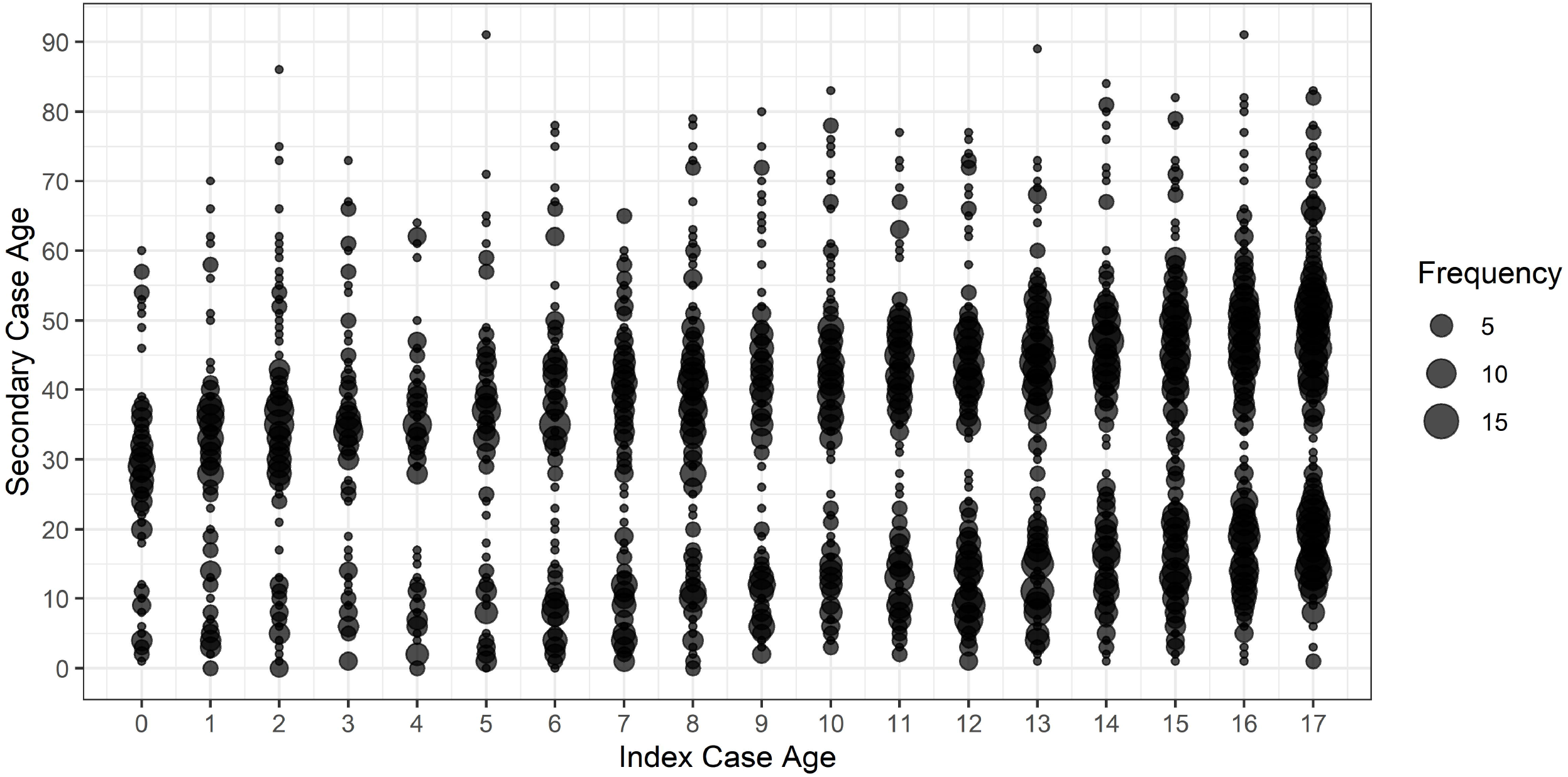
Bubble plot of age-to-age transmission

The proportion of index cases in each age group increased with age, with 12% aged 0-3, 20% aged 4-8, 30% aged 9-13, and 38% aged 14-17 (Table 1). Compared to index cases in the oldest age group, younger index cases had a higher proportion associated with a school/childcare outbreak and shorter testing delays. Index cases aged 4-8 years and 9-13 years had higher proportion with no symptoms reported compared to index cases 14-17 years or index cases 0-3 years. Across all age groups, more index cases had disease onset in the fall/winter (September-December) compared to the summer (June-August), which aligns with the trajectory of the second wave of the pandemic in Ontario.

**Table 1.** Characteristics of pediatric index cases by age group

|  | **0-3 years**<br>**(N=766)** | **4-8 years**<br>**(N=1,257)** | **9-13 years**<br>**(N=1,881)** | **14-17 years**<br>**(N=2,376)** | **>17 years**<br>**(N=82,911)** |
| --- | --- | --- | --- | --- | --- |
| Age, years (median, IQR) | 2 [1, 2] | 6 [5, 7] | 11 [10, 12] | 16 [15, 17] | 40 [28, 54] |
| Gender (N, %) |  |  |  |  |  |
| Female | 374 (49.1) | 586 (46.9) | 820 (43.9) | 1,083 (45.7) | 39,891 (48.3) |
| Male | 385 (50.5) | 663 (53.0) | 1,045 (55.9) | 1,283 (54.1) | 42,572 (51.5) |
| Month of disease onset (N, %) |  |  |  |  |  |
| June | 26 (3.4) | 33 (2.6) | 35 (1.9) | 63 (2.7) | 2,945 (3.6) |
| July | 18 (2.3) | 30 (2.4) | 31 (1.6) | 69 (2.9) | 2,229 (2.7) |
| August | 13 (1.7) | 22 (1.8) | 27 (1.4) | 81 (3.4) | 1,744 (2.1) |
| September | 87 (11.4) | 155 (12.3) | 167 (8.9) | 227 (9.6) | 8,081 (9.7) |
| October | 126 (16.4) | 183 (14.6) | 299 (15.9) | 378 (15.9) | 12,557 (15.1) |
| November | 186 (24.3) | 342 (27.2) | 548 (29.1) | 638 (26.9) | 21,934 (26.5) |
| December | 310 (40.5) | 492 (39.1) | 774 (41.1) | 920 (38.7) | 33,421 (40.3) |
| Testing delay*, days (median, IQR) | 1 [0, 4] | 1 [0, 4] | 2 [0, 4] | 2 [0, 5] | 2 [1, 4] |
| Testing delay distribution* (N, %) |  |  |  |  |  |
| Asymptomatic† | 104 (15.3) | 236 (22.5) | 324 (20.1) | 278 (13.1) | 6,541 (9.1) |
| <0 days‡ | 37 (5.4) | 71 (6.8) | 72 (4.5) | 107 (5.1) | 3,502 (4.9) |
| 0 days | 97 (14.3) | 106 (10.1) | 135 (8.4) | 166 (7.8) | 7,069 (9.8) |
| 1 day | 113 (16.6) | 149 (14.2) | 219 (13.6) | 298 (14.1) | 11,137 (15.5) |
| 2 days | 84 (12.4) | 124 (11.8) | 223 (13.8) | 283 (13.4) | 11,242 (15.6) |
| 3 days | 69 (10.1) | 97 (9.3) | 153 (9.5) | 242 (11.4) | 8,794 (12.2) |
| 4 days | 41 (6.0) | 54 (5.2) | 114 (7.1) | 194 (9.2) | 6,122 (8.5) |
| ≥5 days | 135 (19.9) | 211 (20.1) | 372 (23.1) | 548 (25.9) | 17,619 (24.5) |
| Average family size (median, IQR) | 3.2 [2.9, 3.6] | 3.2 [3.0, 3.5] | 3.3 [3.0, 3.6] | 3.3 [3.1, 3.7] | 3.3 [3.0, 3.6] |
| Symptoms reported (N, %) | 586 (76.5) | 822 (65.4) | 1,296 (68.9) | 1,856 (78.1) | 65,964 (79.6) |
| Associated with a school/childcare outbreak (N, %) | 128 (16.7) | 229 (18.2) | 407 (21.6) | 154 (6.5) | 588 (0.7) |
| Before school/childcare reopening§ (N, %) | 8 (1.0) | 99 (7.9) | 108 (5.7) | 254 (10.7) | 8,189 (9.9) |
| Household size (median, IQR) | 4 [3, 5] | 4 [4, 5] | 4 [4, 5] | 4 [4, 5] | 3 [2, 4] |

\*778 pediatric index cases were excluded from the testing delay models that had no COVID-19 symptoms reported in provincial reportable disease systems, were missing symptom onset date, and were not reported as asymptomatic.
†Index cases that were asymptomatic were identified as cases that were missing symptom onset date (thus specimen collection date was used) and were reported as asymptomatic in provincial reportable disease systems.
‡Index cases with a testing delay of <0 days were those who were tested prior to the onset of their symptoms.
§Reopening dates: June 12, 2020 for index cases aged 0-3 years, September 8, 2020 for index cases aged >3 years and residing outside Toronto, September 15, 2020 for index cases aged >3 years and residing in Toronto.

### Associations with index case characteristics

Compared to index cases aged 14-17 years, index cases aged 0-3 years had higher odds of household transmission in all three models (crude model OR=1.20, 95% CI: 1.01-1.44; adjusted model 1 OR=1.21, 95% CI: 1.01-1.45; adjusted model 2 OR=1.43, 95% CI: 1.17-1.75) (Table 2). There were no significant differences in the odds of transmission for the 4-8 and 9-13 year age groups, with the exception of adjusted model 2 for index cases aged 4-8 years (1.40, 95% CI: 1.18-1.67). Additionally, there were incremental odds of transmission with longer testing delays compared to a 0-day delay (ORs: 1-day delay=1.24, 2-day delay=1.59, 3-day delay=1.97, 4-day delay=2.38, _≥_5-day delay=2.98), as well as increased odds of transmission with larger average family size (1.63 per person increase, 95% CI: 1.43-1.86). No significant differences were observed by gender or by month of disease onset.

**Table 2.** Adjusted odds ratios and 95% confidence intervals for the associations between index case age group and odds of household transmission

|  | **Index cases**<br>**with**<br>**no household**<br>**transmission**<br>**(N, %)** | **Index cases**<br>**with**<br>**household**<br>**transmission**<br>**(N, %)** | **Crude Model**<br>**OR (95% CI)** | **Adjusted**<br>**Model 1\***<br>**OR (95% CI)** | **Adjusted**<br>**Model 2†**<br>**OR (95% CI)** |
| --- | --- | --- | --- | --- | --- |
| Age |  |  |  |  |  |
| 0-3 years | 532 (11.7) | 234 (13.6) | 1.20 (1.01 - 1.44) | 1.21 (1.01 - 1.45) | 1.43 (1.17 - 1.75) |
| 4-8 years | 909 (19.9) | 348 (20.3) | 1.05 (0.90 - 1.22) | 1.06 (0.90 - 1.23) | 1.40 (1.18 - 1.67) |
| 9-13 years | 1,382 (30.3) | 499 (29.1) | 0.99 (0.86 - 1.13) | 0.97 (0.85 - 1.11) | 1.13 (0.97 - 1.32) |
| 14-17 years | 1,740 (38.1) | 636 (37.0) | Ref | Ref | Ref |
| Male gender | 2,433 (53.6) | 943 (55.2) | - | 1.07 (0.95 - 1.19) | 1.09 (0.96 - 1.23) |
| Month of disease onset |  |  |  |  |  |
| June | 119 (2.6) | 38 (2.2) | - | Ref | Ref |
| July | 113 (2.5) | 35 (2.0) | - | 0.98 (0.58 - 1.66) | 1.05 (0.58 - 1.89) |
| August | 112 (2.5) | 31 (1.8) | - | 0.89 (0.52 - 1.52) | 0.93 (0.51 - 1.69) |
| September | 482 (10.6) | 154 (9.0) | - | 1.01 (0.67 - 1.51) | 0.97 (0.62 - 1.51) |
| October | 712 (15.6) | 274 (16.0) | - | 1.22 (0.82 - 1.80) | 1.20 (0.78 - 1.84) |
| November | 1,199 (26.3) | 515 (30.0) | - | 1.37 (0.93 - 2.00) | 1.38 (0.91 - 2.09) |
| December | 1,826 (40.0) | 670 (39.0) | - | 1.16 (0.80 - 1.70) | 1.14 (0.76 - 1.72) |
| Testing delay |  |  |  |  |  |
| Asymptomatic | 853 (18.8) | 89 (5.2) | - | - | 0.34 (0.25 - 0.47) |
| <0 days | 251 (5.5) | 36 (2.1) | - | - | 0.50 (0.33 - 0.75) |
| 0 days | 1,076 (23.8) | 206 (12.1) | - | - | Ref |
| 1 day | 570 (12.6) | 209 (12.3) | - | - | 1.24 (0.95 - 1.61) |
| 2 days | 488 (10.8) | 226 (13.3) | - | - | 1.59 (1.22 - 2.07) |
| 3 days | 361 (8.0) | 200 (11.7) | - | - | 1.97 (1.49 - 2.59) |
| 4 days | 238 (5.3) | 165 (9.7) | - | - | 2.38 (1.77 - 3.19) |
| ≥5 days | 692 (15.3) | 574 (33.7) | - | - | 2.98 (2.34 - 3.80) |
| Average family size | 3.3 [3.0, 3.6] | 3.4 [3.1, 3.7] | - | - | 1.63 (1.43 - 1.86) |

\*Adjusted for gender and month of disease onset.
†Adjusted for gender, month of disease onset, testing delay, and average family size. 778 index cases were excluded from the model that had no COVID-19 symptoms reported in provincial reportable disease systems, were missing symptom onset date, and were not reported as asymptomatic.

In stratified analyses, we did not observe significant heterogeneity (p<0.05) in the odds of household transmission between index cases with versus without symptoms reported, index cases not associated versus associated with a school/childcare outbreak, or index cases with disease onset before versus after school/childcare reopening (Supplementary Tables 1 and 2).

Sensitivity analyses of the crude model and adjusted model 1 resulted in the same direction of association for the 0-3 year age group, but confidence intervals widened (Supplementary Table 3). In adjusted model 2, associations were largely unchanged with the 2-14 day definition, in the symptomatic case analysis, and when controlling for individual-level household size for the 0-3 year age group (adjusted model 2 OR=1.43, 95% CI: 1.17-1.75; 2-14 day OR=1.37, 95% CI 1.11-1.69; symptomatic OR=1.32, 95% CI 1.06-1.64; household size OR=1.31, 95% CI: 1.02-1.67) and 4-8 year age group (adjusted model 2 OR=1.40, 95% CI: 1.18-1.67; 2-14 day OR=1.33, 95% CI 1.11-1.60; symptomatic OR=1.33, 95% CI 1.10-1.61; household size OR=1.31, 95% CI: 1.06-1.62). Associations for the 0-3 year and 4-8 year age groups were also similar to adjusted model 2 in the combined 2-14 day definition and symptomatic case analysis (0-3 year OR=1.25, 95% CI 1.00-1.57; 4-8 year OR=1.23, 95% CI 1.00-1.50), as well as for the 0-3 year age group with the 4-14 day definition (1.35, 95% CI 1.07-1.70) and after further restricting to symptomatic cases (1.26, 95% CI 0.96-1.64).

## Discussion

In this study of 6,280 pediatric index cases, we observed that children aged 0-3 years had greater odds of household transmission compared to children aged 14-17 years. This association was observed irrespective of factors such as presence of symptoms, school/childcare reopening, or association with a school/childcare outbreak. We also observed some evidence of greater odds of household transmission for children aged 4-8 years after controlling for testing delays and neighbourhood-level average family size (as well as individual-level household size). We identified clustering of ages in our age-to-age transmission plot, which likely reflects the age structure of households with younger pediatric cases living with, and transmitting to, younger caregivers and siblings.

To date, there have been challenges with comparing the roles of children in household spread of SARS-CoV-2 due to small numbers of pediatric cases. Zhu et al.^2^ and Thompson et al.^15^ conducted meta-analyses of studies examining the age of index cases in households, and found that 3%-19% of households had pediatric index cases, depending on the definitions of index cases and pediatric cases (<18 years and <20 years, respectively). We identified pediatric index cases aged <18 years in approximately 7% of all households (6,280/89,191 households), which is similar to studies from Greece (9% <18 years)^26^, Switzerland (8% <16 years)^27^, Denmark (5% <20 years)^21^, Hunan, China (5% <15 years)^19^, and Guangzhou, China (5% <20 years)^9^; but higher than studies from South Korea (3% <20 years)^20^ and Wuhan, China (1% <20 years)^12^, and lower than a study from the USA (14% <18 years).^23^ Presumed household transmission occurred in 27% of households with a pediatric index case in our study using the 1-14 day definition of secondary transmission (24% and 16% using 2-14 and 4-14 day definitions, respectively), which is close to an estimate from Catalonia, Spain (28%).^16^

A small number of studies have compared associations between age of pediatric index cases and household transmission with mixed findings. Our results align with Danish studies from Lyngse et al.^21,22^, which found that among children there were increased odds of transmission with younger age. In the Danish study the OR for children aged 0-5 was 1.11 (95% CI: 1.01-1.19) compared to the 30-35 year reference group, versus 0.82 (95% CI: 0.78-0.85) for children aged 10-15.^22^ Soriano-Arandes et al.^16^ also found that the highest OR for transmission was among index cases 0-2 years (2.27, 95% CI: 0.62-8.35 versus 12-15 year group), but confidence intervals were wide.

Grijalva et al.^23^ observed high household secondary attack rates (SAR) for both pediatric and adult index cases in Tennessee and Wisconsin, USA; the SAR for index cases <12 years was 53% (95% CI: 31%-74%) and for cases 12-17 years was 38% (95% CI: 23%-56%). This compared to 55% (95% CI: 46%-64%) for cases 18-49 years and 62% (95% CI: 44%-77%) for cases _≥_50 years. Thompson et al.^15^ conducted a meta-analysis of studies and reported no significant difference in household SAR for index cases 0-9 years versus index cases 10-19 years. Conversely, Park et al.^20^ found that household contacts of index cases 10-19 years had the highest SAR among 10-year age groups of both children and adults at 18.6% (95% CI 14.0%-24.0%), compared to 5.3% (95% CI 1.3%-13.7%) for index cases 0-9 years.

Differences in viral shedding, symptom expression, and behavioral factors may explain differences in the odds of household transmission for pediatric age groups across studies.^4,5,21,22^ Viral load is suspected to be an important factor affecting the odds of SARS-CoV-2 transmission.^22,28,29^ Several studies of age-specific viral shedding of SARS-CoV-2 have reported that viral loads in children are similar or higher than viral loads in adults.^22,30–33^ In particular, one study reported that children <5 years of age might carry more viral RNA in their nasopharynx than older children and adults.^30^ Additionally, biases in testing practices, such as the preferential testing of symptomatic cases and contacts, inherently leads to under-detection of pediatric cases and challenges estimating their rates of transmission.^4,5,15^ Studies have found younger children are more likely to be asymptomatic^3,16,26^, which has been postulated as a reason for lower infectivity as lower SARs have been reported for asymptomatic index cases compared to symptomatic index cases.^2,12,15,24^ We found that asymptomatic status and testing delays had strong gradient effects on transmission, similar to our previous study of household transmission.^25^ However, even after adjusting for the lower odds of asymptomatic transmission and testing delays in our study, children 0-3 years and 4-8 years remained associated with higher household transmission than children 14-17 years. A possible explanation for this finding, as mentioned by Lyngse et al.^21^ and supported by our findings, is that younger children are not able to self-isolate from their caregivers when they are sick, irrespective of the timing of testing. Finally, Li et al.^12^ observed both increased risk of transmission from individuals <20 years and faster isolation of pediatric cases in Wuhan, suggesting possible higher infectivity of children compared to adults.

This study has some limitations that should be acknowledged. First, there is the possibility of misclassifying household transmission if secondary case infection was truly acquired outside of the household, or if the true index case of the household was untested. This is particularly relevant for pediatric cases due to their increased probability of having mild or asymptomatic infection, and thus increased probability of infection being missed. We attempted to account for this in sensitivity analyses by modifying the secondary case definition from 1-14 days to 2-14 and 4-14 days, and restricting analysis to symptomatic cases only. Second, this study used multiple evolving data systems for reporting COVID-19 cases in Ontario. As a result, there was some inconsistency regarding how symptoms were reported, which may result in some misclassification of symptomatic cases. We carefully examined the reporting practices over the months covered in the study, and selected a combination of variables (symptoms reported, symptom onset date available, and/or asymptomatic flag) we felt reflected the most likely symptom status of cases. Third, we could not reliably calculate SARs in the study as we did not know the number of non-infected contacts in households for the full cohort. However, after controlling for individual-level household size within the subset of the cohort with this information available, our conclusions were unaltered.

This study also has several strengths. This is a large, population-based study of all individuals with confirmed SARS-CoV-2 infection in Canada’s most populated province, thus we had sufficient data to explore transmission within the understudied pediatric group. We were also able to include relevant covariates such as testing delays and household size; our results suggest that early testing of pediatric index cases and reduced household size/crowding may be useful strategies to minimize secondary household transmission. Second, the use of a natural language processing algorithm to perform address matching allowed us to reliably identify cases in the same household, rather than relying on contact tracing or other types of epidemiological linkage that would be difficult to perform for high volumes of cases. Third, our application of various sensitivity analyses for symptom status and the definition of secondary transmission increased our certainty of the direction of transmission from index case to secondary case, further supporting our findings.

## Conclusion

As the number of pediatric cases increases worldwide, the role of children in household transmission will continue to grow. We found that younger children are more likely to transmit SARS-CoV-2 infection compared to older children, and the highest odds of transmission were observed for children aged 0-3 years. Differential infectivity of pediatric age groups has implications for infection prevention controls within households and schools/childcare to minimize risk of household secondary transmission. Although children do not appear to transmit infection as frequently as adults, caregivers should be aware of the risk of transmission while caring for sick children in the household setting. As it is challenging and often impossible to socially isolate from sick children, caregivers should apply other infection control measures where feasible, such as use of masks, increased hand washing, and separation from siblings.

## Data Availability

Data sharing requests should be directed to Public Health Ontario.

## Acknowledgements

The authors thank James Johnson and Arezou Saedi for conducting the address-matching work, Trevor van Ingen for providing the neighbourhood-level data, and Semra Tibebu for cleaning the individual-level household size variable.

## Contributors

LP performed the analysis and drafted the manuscript. SB, ND, KS, MS, and KB conceptualized the study. SB, ND, KB, KS, MS, MW, and LP developed the methodology. SB, KB, and LP verified the underlying data. SB, ND, KS, MS, KB, MW, and EC reviewed the manuscript.

## Declaration of interests

The authors have no conflicts of interest to declare.

## Data sharing

Data sharing requests should be directed to Public Health Ontario.

## Notes

### Competing Interest Statement

The authors have declared no competing interest.

### Funding Statement

This study was supported by Public Health Ontario.

### Author Declarations

We obtained ethics approval from Public Health Ontario's Research Ethics Board.

