## Supplementary Material for "Pediatric household transmission of SARS-CoV-2 infection"

**Supplementary Table 1.** Stratified analyses for the associations between index case age group and odds of household transmission

|  | **Index cases with**  **no household transmission**  **(N, %)** | **Index cases with**  **household transmission**  **(N, %)** | **Crude Model**  **OR (95% CI)** | **Adjusted  Model 1***  **OR (95% CI)** | **Adjusted Model 2†**  **OR (95% CI)** |
| --- | --- | --- | --- | --- | --- |
| Asymptomatic‡ |  |  |  |  |  |
| 0-3 years | 156 (10.1) | 24 (13.4) | 1.62 (0.96 - 2.75) | 1.64 (0.96 - 2.80) | 1.49 (0.68 - 3.26) |
| 4-8 years | 383 (24.9) | 52 (29.1) | 1.43 (0.94 - 2.18) | 1.41 (0.92 - 2.16) | 1.31 (0.70 - 2.45) |
| 9-13 years | 527 (34.2) | 58 (32.4) | 1.16 (0.77 - 1.75) | 1.13 (0.75 - 1.71) | 1.20 (0.66 - 2.18) |
| 14-17 years | 475 (30.8) | 45 (25.1) | Ref | Ref | Ref |
| Symptomatic |  |  |  |  |  |
| 0-3 years | 376 (12.4) | 210 (13.7) | 1.20 (0.98 - 1.45) | 1.20 (0.99 - 1.46) | 1.41 (1.15 - 1.74) |
| 4-8 years | 526 (17.4) | 296 (19.2) | 1.20 (1.01 - 1.43) | 1.22 (1.03 - 1.46) | 1.41 (1.17 - 1.69) |
| 9-13 years | 855 (28.3) | 441 (28.7) | 1.10 (0.95 - 1.28) | 1.09 (0.93 - 1.26) | 1.12 (0.96 - 1.31) |
| 14-17 years | 1,265 (41.9) | 591 (38.4) | Ref | Ref | Ref |
| No association with school/childcare outbreak |  |  |  |  |  |
| 0-3 years | 433 (11.1) | 205 (13.9) | 1.29 (1.07 - 1.56) | 1.30 (1.07 - 1.57) | 1.48 (1.19 - 1.83) |
| 4-8 years | 745 (19.2) | 283 (19.1) | 1.04 (0.88 - 1.22) | 1.05 (0.88 - 1.23) | 1.36 (1.12 - 1.64) |
| 9-13 years | 1,080 (27.8) | 394 (26.7) | 1.00 (0.86 - 1.15) | 0.99 (0.85 - 1.15) | 1.09 (0.92 - 1.29) |
| 14-17 years | 1,626 (41.9) | 596 (40.3) | Ref | Ref | Ref |
| Association with school/childcare outbreak |  |  |  |  |  |
| 0-3 years | 99 (14.6) | 29 (12.1) | 0.83 (0.48 - 1.45) | 0.95 (0.54 - 1.67) | 1.26 (0.65 - 2.46) |
| 4-8 years | 164 (24.2) | 65 (27.2) | 1.13 (0.71 - 1.79) | 1.16 (0.73 - 1.85) | 1.54 (0.87 - 2.70) |
| 9-13 years | 302 (44.5) | 105 (43.9) | 0.99 (0.65 - 1.51) | 0.99 (0.64 - 1.51) | 1.26 (0.76 - 2.08) |
| 14-17 years | 114 (16.8) | 40 (16.7) | Ref | Ref | Ref |
| Before school/childcare reopening§ |  |  |  |  |  |
| 0-3 years | 4 (1.1) | 4 (3.4) | 3.16 (0.77 - 13.03) | 2.98 (0.69 - 12.83) | 5.41 (1.02 - 28.66) |
| 4-8 years | 77 (21.9) | 22 (18.8) | 0.90 (0.52 - 1.57) | 0.90 (0.51 - 1.57) | 1.09 (0.58 - 2.06) |
| 9-13 years | 78 (22.2) | 30 (25.6) | 1.22 (0.73 - 2.03) | 1.19 (0.72 – 2.00) | 1.16 (0.64 - 2.09) |
| 14-17 years | 193 (54.8) | 61 (52.1) | Ref | Ref | Ref |
| After school/childcare reopening |  |  |  |  |  |
| 0-3 years | 528 (12.5) | 230 (14.4) | 1.17 (0.98 - 1.41) | 1.27 (1.05 - 1.52) | 1.49 (1.21 - 1.83) |
| 4-8 years | 832 (19.8) | 326 (20.4) | 1.05 (0.90 - 1.24) | 1.07 (0.91 - 1.26) | 1.43 (1.19 - 1.72) |
| 9-13 years | 1,304 (31.0) | 469 (29.3) | 0.97 (0.84 - 1.12) | 0.96 (0.83 - 1.11) | 1.13 (0.97 - 1.33) |
| 14-17 years | 1,547 (36.7) | 575 (35.9) | Ref | Ref | Ref |

*Adjusted for gender and month of disease onset.
†Adjusted for gender, month of disease onset, testing delay, and average family size. 778 index cases were excluded from the model that had no COVID-19 symptoms reported in provincial reportable disease systems, were missing symptom onset date, and were not reported as asymptomatic. The “no symptoms reported” stratified analysis did not include testing delay in adjusted model 2 (and thus no case exclusion was applied), as by definition these cases were all classified in the asymptomatic group of the testing delay variable.
‡Index cases with no symptoms reported were defined as cases that were missing symptom onset date (thus specimen collection date was used) and were reported as asymptomatic in provincial reportable disease systems.
§Reopening dates: June 12, 2020 for index cases aged 0-3 years, September 8, 2020 for index cases aged >3 years and residing outside Toronto, September 15, 2020 for index cases aged >3 years and residing in Toronto.

**Supplementary Table 2.** Heterogeneity p-values corresponding to stratified analyses

|  | **Crude Model** | **Adjusted Model 1** | **Adjusted Model 2** |
| --- | --- | --- | --- |
|  | 0-3 years | | |
| Symptoms reported | 0.37 | 0.36 | 0.91 |
| School/childcare outbreak | 0.10 | 0.27 | 0.65 |
| School/childcare reopening | 0.53 | 0.58 | 0.58 |
|  | 4-8 years | | |
| Symptoms reported | 0.49 | 0.57 | 0.83 |
| School/childcare outbreak | 0.76 | 0.71 | 0.71 |
| School/childcare reopening | 0.59 | 0.55 | 0.40 |
|  | 9-13 years | | |
| Symptoms reported | 0.82 | 0.88 | 0.84 |
| School/childcare outbreak | 0.97 | 1.00 | 0.63 |
| School/childcare reopening | 0.46 | 0.49 | 0.94 |

**Supplementary Table 3.** Sensitivity analysis for the associations between index case age group and odds of household transmission

|  | **Index cases with**  **no household transmission**  **(N, %)** | **Index cases with**  **household transmission**  **(N, %)** | **Crude Model**  **OR (95% CI)** | **Adjusted Model 1***  **OR (95% CI)** | **Adjusted Model 2†**  **OR (95% CI)** |
| --- | --- | --- | --- | --- | --- |
| Secondary cases occurring 2-14 days after index case |  |  |  |  |  |
| 0-3 years | 562 (11.8) | 204 (13.6) | 1.15 (0.95 - 1.38) | 1.15 (0.96 - 1.39) | 1.37 (1.11 - 1.69) |
| 4-8 years | 959 (20.1) | 298 (19.9) | 0.98 (0.84 - 1.15) | 0.99 (0.84 - 1.16) | 1.33 (1.11 - 1.60) |
| 9-13 years | 1,456 (30.4) | 425 (28.4) | 0.92 (0.80 - 1.06) | 0.90 (0.78 - 1.04) | 1.04 (0.88 - 1.22) |
| 14-17 years | 1,805 (37.7) | 571 (38.1) | Ref | Ref | Ref |
| Secondary cases occurring 4-14 days after index case |  |  |  |  |  |
| 0-3 years | 624 (11.9) | 142 (13.9) | 1.11 (0.90 - 1.37) | 1.12 (0.91 - 1.39) | 1.35 (1.07 - 1.70) |
| 4-8 years | 1,073 (20.4) | 184 (18.0) | 0.84 (0.69 - 1.01) | 0.84 (0.69 - 1.01) | 1.10 (0.90 - 1.36) |
| 9-13 years | 1,586 (30.2) | 295 (28.8) | 0.91 (0.77 - 1.07) | 0.88 (0.75 - 1.04) | 1.04 (0.87 - 1.25) |
| 14-17 years | 1,972 (37.5) | 404 (39.4) | Ref | Ref | Ref |
| Control for household size‡ |  |  |  |  |  |
| 0-3 years | 311 (11.9) | 145 (13.0) | - | - | 1.31 (1.02 - 1.67) |
| 4-8 years | 531 (20.3) | 229 (20.5) | - | - | 1.31 (1.06 - 1.62) |
| 9-13 years | 794 (30.3) | 318 (28.4) | - | - | 1.07 (0.89 - 1.29) |
| 14-17 years | 986 (37.6) | 427 (38.2) | - | - | Ref |
| Cohort restricting to symptomatic cases |  |  |  |  |  |
| 0-3 years | 434 (12.7) | 176 (13.5) | 1.12 (0.91 - 1.37) | 1.13 (0.92 - 1.38) | 1.32 (1.06 - 1.64) |
| 4-8 years | 605 (17.7) | 250 (19.1) | 1.14 (0.95 - 1.36) | 1.16 (0.97 - 1.39) | 1.33 (1.10 - 1.61) |
| 9-13 years | 976 (28.5) | 370 (28.3) | 1.04 (0.89 - 1.22) | 1.03 (0.88 - 1.20) | 1.06 (0.90 - 1.25) |
| 14-17 years | 1,411 (41.2) | 512 (39.1) | Ref | Ref | Ref |
| Secondary cases occurring 2-14 days after index case in cohort restricting to symptomatic cases |  |  |  |  |  |
| 0-3 years | 464 (12.8) | 146 (13.3) | 1.06 (0.86 - 1.31) | 1.06 (0.86 - 1.32) | 1.25 (1.00 - 1.57) |
| 4-8 years | 651 (17.9) | 204 (18.6) | 1.06 (0.87 - 1.28) | 1.06 (0.88 - 1.29) | 1.23 (1 .00- 1.50) |
| 9-13 years | 1,037 (28.5) | 309 (28.1) | 1.00 (0.85 - 1.19) | 0.98 (0.83 - 1.16) | 1.01 (0.85 - 1.20) |
| 14-17 years | 1,483 (40.8) | 440 (40.0) | Ref | Ref | Ref |
| Secondary cases occurring 4-14 days after index case in cohort restricting to symptomatic cases |  |  |  |  |  |
| 0-3 years | 515 (12.8) | 95 (13.5) | 1.06 (0.82 - 1.36) | 1.07 (0.83 - 1.38) | 1.26 (0.96 - 1.64) |
| 4-8 years | 733 (18.2) | 122 (17.3) | 0.95 (0.76 - 1.20) | 0.96 (0.76 - 1.21) | 1.11 (0.87 - 1.41) |
| 9-13 years | 1,145 (28.4) | 201 (28.6) | 1.00 (0.83 - 1.22) | 0.98 (0.80 - 1.19) | 1.00 (0.81 - 1.23) |
| 14-17 years | 1,637 (40.6) | 286 (40.6) | Ref | Ref | Ref |

*Adjusted for gender and month of disease onset.
†Adjusted for gender, month of disease onset, testing delay, and average family size. 778 index cases were excluded from the model that had no COVID-19 symptoms reported in provincial reportable disease systems and were not reported as asymptomatic.
‡Controlled for household size instead of average family size. Analysis included 3,741/6,280 (59.6%) pediatric index cases with household size information available.
